# NURSE-led care in Patients Undergoing CATheter Ablation for Atrial Fibrillation: The NURSECAT-AF randomized trial

**DOI:** 10.1101/2025.06.29.25330023

**Authors:** Alba Cano-Valls, Maria-Antonia Martinez-Monblan, Esther Carro-Fernández, Mireia Niebla-Bellido, Rebeca Domingo-Criado, Sara Hevia-Puyo, Montserrat Venturas-Nieto, Roger Borras, José María Tolosana, Andreu Porta-Sánchez, Jean-Baptiste Guichard, Till Althoff, Ivo Roca-Luque, Lluis Mont, Eduard Guasch

**Affiliations:** Nursing faculty, Universitat de Barcelona, Barcelona, Catalonia, Spain; Cardiology department, Clinic Barcelona, Barcelona, Catalonia, Spain; Institut d’Investigacions Biomédiques August Pi I Sunyer (IDIBAPS), Barcelona, Catalonia, Spain; Centro de Investigación Biomédica en Red-SAM (CIBERSAM); Medicine faculty, Universitat de Barcelona, Barcelona, Catalonia, Spain; Centro de Investigación Biomédica en Red-CV (CIBERCV), Spain

**Author notes:** To whom correspondence should be sent: Maria Antonia Martínez Monblán, Carrer de la Feixa Llarga, s/n, 08907 L’Hospitalet de Llobregat, Barcelona, Spain Tel: +34. 934 02 42 92. Share senior authorship (Lluis Mont, Eduard Guasch).

**Keywords:** Atrial fibrillation, Ablation, Randomized trial, Education, Nurse-led management, Integrated care

## Abstract

**Background:** Atrial fibrillation (AF) is associated with reduced quality of life and frequent hospitalizations. Integrated nurse-led care has proven beneficial in unselected AF patients, but evidence specific to patients undergoing catheter ablation is limited. We aimed to assess the impact of a structured nurse-led intervention in patients undergoing first-time AF ablation.

**Methods:** NURSECAT-AF was a single-center prospective randomized clinical trial comparing usual care (UC) with a nurse-led peri-ablation care (NLC) which incorporated an educational program on AF, peri-procedural support, and risk factor management. Consecutive patients without heart failure referred for first-time AF ablation were randomized to UC or NLC. Visits in NLC were scheduled at 15 days pre-ablation, and 15 days, 3 months and 6 months post-ablation. The primary endpoint was quality of life at 12 months post-ablation using the Arrhythmia-Specific Scale in Tachycardia and Arrhythmia (ASTA). Secondary outcomes included arrhythmia recurrence, readmissions and emergency visits, and symptom burden at one year, and AF knowledge and satisfaction at 3 months.

**Results:** Of 116 patients screened, 66 were randomized (33 per group; mean age 63±10 years; 67% male). At 12 months, the NLC group showed significantly better quality of life (baseline-adjusted ASTA difference +4 points [95%CI 1.8-6.3], p=0.007) than UC, and presented with less arrhythmia recurrences (OR 0.2 [95%CI 0.05-0.78]) and emergency visits (OR 0.2 [95%CI 0.06-0.66]). Patients assigned to NLC also presented with a lower symptom burden, higher satisfaction and greater disease knowledge. Risk factor profile was improved in the NLC group, with higher rates of smoking cessation, engagement in regular physical activity, and weight optimization. Nurse-led management enabled more frequently diagnosing obstructive sleep apnea.

**Conclusion:** Nurse-led, integrated care for patients undergoing AF ablation improves the quality of life, clinical outcomes and risk factor management at one year post-procedure. These findings support the incorporation of structured nurse-led interventions in the peri-ablation care pathway.

**Clinical Trial Registration:** NCT05333445.

https://clinicaltrials.gov/study/NCT05333445?term=NCT05333445&rank=1

**GRAPHICAL ABSTRACT:** (Annex B)

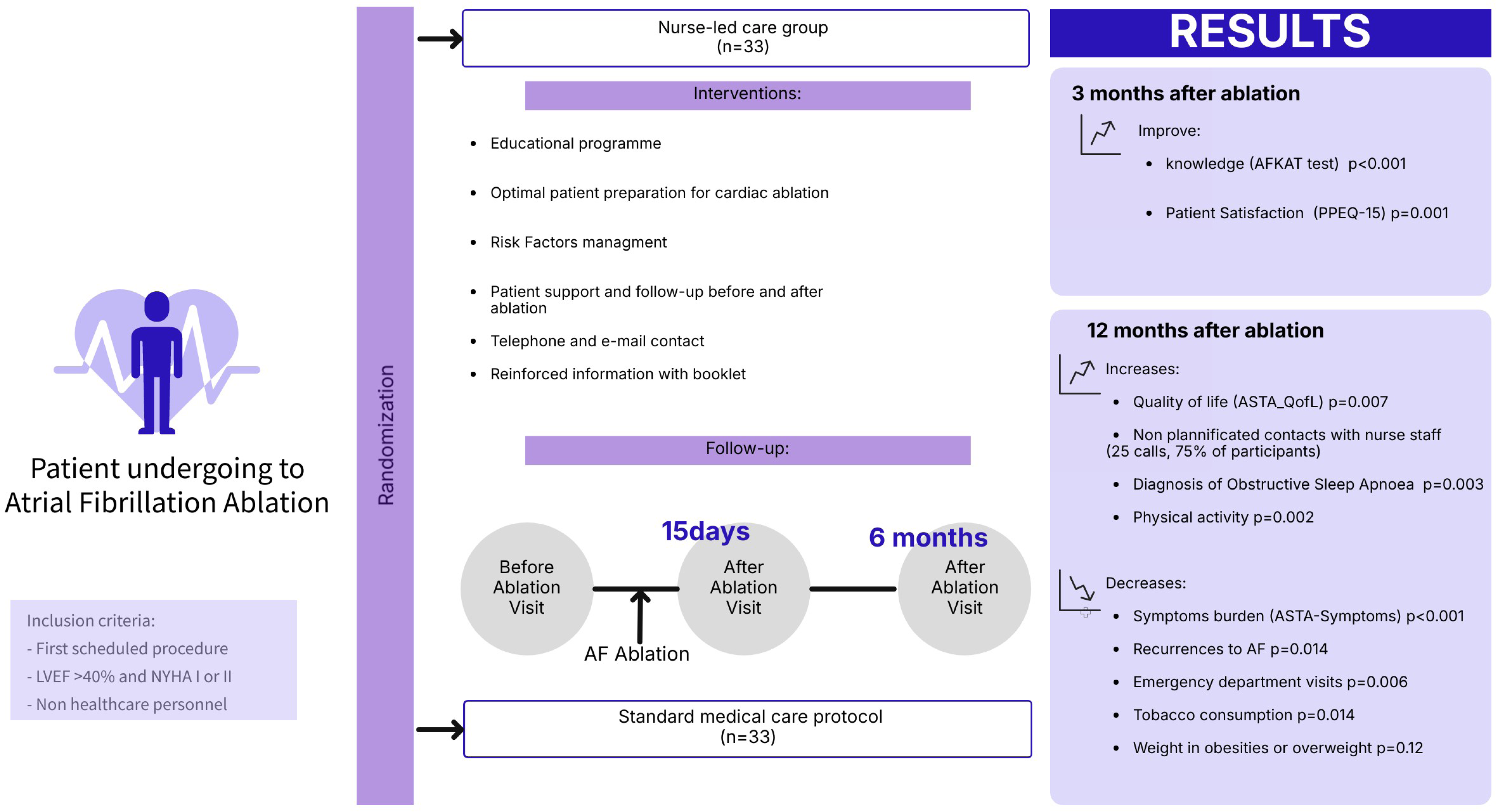

**Clinical Perspective:** *What is known?:*

- Nurse-coordinated, integrated care pathways in patients with chronic conditions, including atrial fibrillation (AF), have been shown to improve patient-reported quality of life and to reduce all-cause and cardiovascular mortality.
- Atrial fibrillation ablation effectively decreases arrhythmic burden and enhances quality of life; nonetheless, recurrences remain common, and many patients continue to experience symptoms and diminished functional status.
- The role of periprocedural nurse-led management remains unknown.

*What this study adds?:*

- In a randomized controlled trial, implementation of a periprocedural, nurse-led care model significantly improved quality of life in patients undergoing AF ablation, as assessed by validated symptom and well-being instruments.
- At one year post-ablation, the incidence of AF recurrence is reduced in the cohort receiving nurse-led care, an effect likely mediated by superior optimization of cardiovascular risk factors achieved through targeted nurse intervention.
- A comprehensive, nurse-delivered strategy—comprising patient education, systematic preparation for ablation, and proactive management of modifiable arrhythmia risk factors—confers meaningful clinical benefit in the management of AF.

## Introduction

The management of atrial fibrillation (AF) has shifted in recent years towards an integrated, multidisciplinary care. Recent studies demonstrate that integrated management of patients with AF positively impacts quality of life, satisfaction, the management of risk factors, treatment adherence, and AF symptom burden. ^1,2^ The role of a nurse practitioner is central to the success of this approach. Nurse-led education, counselling, and interventions have proven to be effective tools for improving quality of life (QoL), reducing anxiety, and improving outcomes in patients with chronic conditions including heart failure or ischemic heart disease. ^3-4^ Emerging data support the role of nursing education and counselling for AF patients in the outpatient setting, and more recently as part of an integrated multidisciplinary approach that also involves hospitalization.^5–7^

Catheter ablation is a cornerstone of the management of many patients with AF in whom sinus rhythm maintenance is sought, providing superior efficacy and improvement in QoL compared with antiarrhythmic drugs.^8^ The efficacy of ablation in the short and long term is closely related to the control of AF risk factors.^9,10^ Before and after ablation patients commonly have multiple contacts with health care professionals, evolving a sweet spot to impact on patient QoL and lifestyle. However, the impact of a nurse-led intervention in the AF ablation periprocedural period remains to be elucidated.^11^

The aim of the NURSECAT-AF trial was to test the effectiveness of expert nurse-led care within an integrated management programme for patients undergoing AF ablation.

## Methods

The NURSECAT-AF was a single-center, prospective, randomized controlled trial designed to compare nurse-led care (NLC) vs usual care (Control) for patients undergoing AF ablation. NURSECAT-AF was registered at clinicaltrials.gov (NCT05333445) before the first patient had been randomized. It was approved by the institutional ethics committee (HCB/2021/1045) and all participants provided written informed consent. The overall study design is depicted in **Figure 1**. The underlying data for this article are available from the corresponding author upon reasonable request.

**Figure 1.**
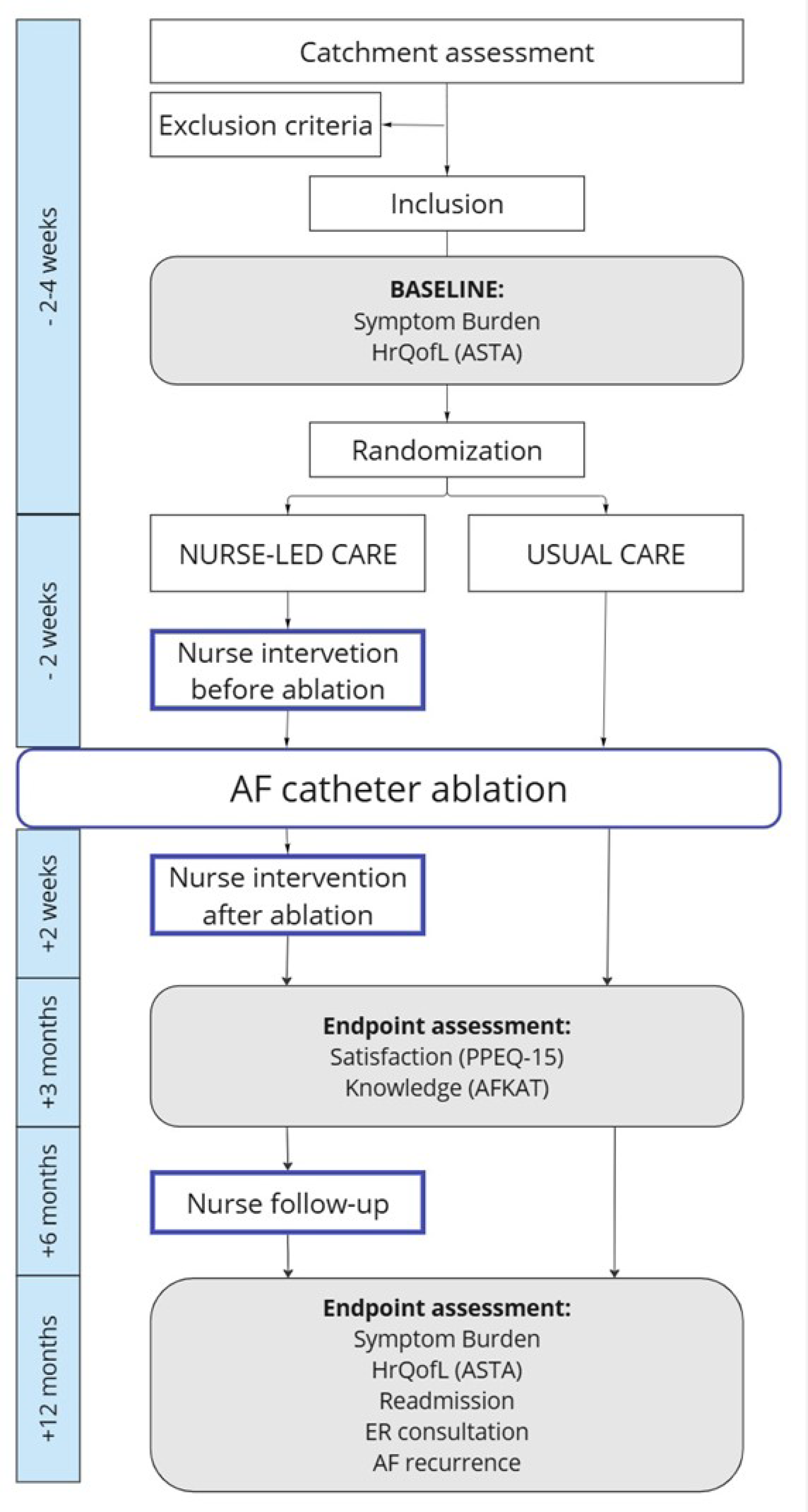
Workflow of the study design.

### Objectives

The primary objective on NURSECAT-AF was to analyse the effect of NLC on QoL assessed with the ASTA questionnaire 12 months after ablation.

Pre-planned secondary objectives (detailed description in the **Supplemental table 1**) were to evaluate the effect of NLC at the 12-month follow-up on the diagnosis and control of AF risk factors; on readmissions and emergency room visits; on the recurrences of AF; and on the burden of AF symptoms. At 3 months, secondary objectives were to assess the level of patient satisfaction with the AF follow-up care process; and to evaluate patient knowledge of AF management. Unplanned contacts received by the AF nurse practitioner via emails or phone calls from study participants were collected during the 12-month follow-up period.

### Study population

Consecutive patients undergoing their first elective catheter ablation for drug-refractory AF at Clínic Barcelona were eligible to participate in the study. Key exclusion criteria were being a healthcare professional, participating in another interfering study, or having a diagnosis of significant heart failure (defined as NYHA class III or higher, or left ventricular ejection fraction [LVEF] less than 40%). The full list of inclusion and exclusion criteria is reported in the **Supplemental table 2**.

Patients were randomized prior to ablation to receive either a nurse intervention plus standard medical care (NLC group) or standard medical care alone (Control group) as summarized in **Figure 1**. Patients were allocated to each group using a predefined randomization list provided by a statistician. The interventional electrophysiologist and follow-up physicians were blinded to the study arm throughout all phases of the study.

Ablation procedures for AF were conducted using standard protocols, under general anaesthesia, and utilizing a three-dimensional mapping system (CARTO, EnSite, or RYTHMYA) or cryoablation, as per institutional guidelines. The ablation strategy involved pulmonary vein isolation for all patients, with additional ablation lines left to the discretion of the treating physician. Following hospital discharge, all patients received follow-up evaluations by their general practitioner and an electrophysiologist at 3, 6 and 12 months, as per standard follow-up. An ECG and a 24-hour ECG-Holter were performed at every visit, and patients were encouraged to obtain an ECG in the presence of symptoms suggestive of arrhythmia.

### Nurse intervention

There was no interaction with the nurse practitioner in the Control arm and all education provided was left to their treating cardiologist and electrophysiologist, consistent with routine care at our center.

The nurse intervention in the NLC group was based on three main components:

1. An educational plan for managing AF,
2. Screening for risk factors with appropriate management, and
3. Preparing the patient before and after the ablation to optimise outcomes (including review of complementary tests, early detection of complications and therapeutic adherence).

These were conducted by a team of nurse practitioners trained in AF management and with extensive experience in the electrophysiology lab. The intervention was implemented at tree points: face-to-face between 15 and 3 days before admission, between 7 and 15 days after ablation, 6 months after the ablation procedure.

During the first meeting, the education was conducted in a structured manner, following the Bowyer model^11^ and using these pre-specified subtitles: "How the Heart Works"; "Causes and Risk Factors of AF"; "Symptoms of AF"; "Treatment Goals in AF"; "Preparation for Ablation Procedure"; and "Lifestyle Modification". The technique involves discussing these main subtitles and adapting the visit to meet each patient’s needs (**Supplemental table 3**). To reinforce the knowledge and information imparted, the research team devised a booklet, which was distributed to all patients in the NLC groups (provided as a **Supplemental material**). Patients were also provided with a telephone number and email address to contact the nurse for any non-urgent questions or concerns throughout the twelve-month follow-up period.

The visits after ablation reviewed possible complications after ablation. In addition, messages from the first visit were reinforced and the focus was on the management of risk factors.

The risk factors were monitored at all nurse visits, emphasizing the importance of their management for proper AF and symptom control. Data were collected through physical examinations, self-completed questionnaires, and medical record reviews. The risk factors evaluated were those recommended by the guidelines.^12^ Diabetes, dyslipidemia, hypertension, obesity, obstructive sleep apnea (OSA), sedentary lifestyle, toxic habits such as smoking or alcohol consumption, and monitoring of thyroid and renal function were evaluated. A blood test was conducted for those patients who did not have one within the past year. Patients with a BMI over 30 were referred to the endocrinology unit. Patients who tested positive on the Berlin questionnaire^13,14^ underwent a polysomnography (PSG). Patients received counselling on strategies to manage specific risk factors and were referred to specialists as needed on a case-by-case basis. Additionally, patient questions and concerns were addressed throughout all four phases of the intervention. In addition, patients could contact the nurse by mail or phone call (available hours 8am-4pm from Monday to Friday).

### Study endpoints

#### Quality of life

The primary outcome was assessed in both groups by measuring at 12 months the QoL domain of the Arrhythmia-Specific Scale in Tachycardia and Arrhythmia (ASTA-QoL) questionnaire, translated and validated into Spanish. ^15,16,17^ The QoL questionnaire is sensitive to AF patients and was self-administered by participants. It consists of 13 questions, with responses measured on a Likert frequency scale from "Never = 0" to "Always = 3". Higher scores on this scale indicate poorer quality of life.

QoL was also assessed with the validated Cantril Ladder tool. Patients self-assessed their quality of life by rating it on a scale from 0 (worst possible quality of life) to 100 (best possible quality).

#### Clinical follow-up

All participants were scheduled for a 12-month follow-up to collect data on the number of any cause hospital readmissions, visits to the emergency department, and documented AF recurrences. Health records from the responsible electrophysiologist were consulted, and the recurrences were reviewed to ensure they had been documented with an ECG.

#### Risk factor management

Modifiable risk factors were assessed at baseline and at 12 months to identify differences between both groups. Body weight and height were measured and the body mass index (BMI) calculated [weight in kg / (stature in m)^2^]. The prevalence of severe OSA diagnosed with a PSG was recorded. Current smokers were identified as those who smoke at least 1 cigarette in the last 30 days. Daily drinking was defined as those patients regularly drinking at least 1 alcohol drink per day. Physical activity was assessed by estimating energy expenditure over 14 days (METS-min/14 days) using the reduced Spanish validated version of the Minnesota Leisure Time Physical Activity Questionnaire (VREM).^18^

#### AF symptoms

The burden of AF symptoms was determined using the ASTA-Symptoms questionnaire and compared at twelve months of follow-up between the two groups. The ASTA symptom questionnaire uses a Likert scale to evaluate 9 types of symptoms, with possible responses ranging from 0 to 3: “No” (0); “Yes, to a certain extent” (1); “Yes, quite a lot” (2); or “Yes, a lot” (3). A summarized score is calculated with data from all items ranging from 0 to 27. A higher score implies higher symptom burden due to the heart rhythm disturbance.

The validated modified EHRA scale which stratifies the impact of experienced symptoms,^19^ was also collected by the nurse practitioner. It consists of 5 levels: asymptomatic (1), mild symptoms that do not affect daily activities (2a), symptoms that are bothersome to the patient but do not interfere with daily activities (2b), symptoms that affect daily activities of the patient (3), and incapacitating symptoms that interrupt daily activities (4).

#### Patient satisfaction

Satisfaction with the medical procedure was assessed at 3 months post-ablation using the validated Spanish Picker Patient experiences-15 questionnaire (PPEQ-15) adapted to our service. ^20^ It consists of 14 closed-ended questions and is conducted using a checklist to assign numerical values to subjective satisfaction using a Likert scale from "No = 0" to "Yes, always = 3". Higher scores on this scale represent greater satisfaction. We show the questions adapted to the AF Ablation procedure in **Supplemental table 4**.

#### Knowledge on AF

Knowledge about AF and its integrated management was explored at 3 months with the Atrial Fibrillation Knowledge Assessment Tool (AFKAT) questionnaire adapted and translated into Spanish by our team. ^21^ The AFKAT questionnaire consists of 25 True/False questions, and higher scores correlate with greater knowledge. The questionnaire was delivered before the medical visit.

#### Unplanned contacts

The number and topic of unplanned emails and phone calls received from participants in the intervention group during the 12 months following ablation was also recorded.

### Statistical Analyses

Baseline qualitative variables are reported as absolute and relative frequencies for each category. Quantitative variables are described using mean and standard deviation (mean±SD) or median (Q1-Q3), depending on whether the variable followed a normal distribution (QQ plots). The 12-month follow-up data were compared using analysis of covariance (ANCOVA). This analysis used the respective initial scores as covariates. ANCOVA adjusts for any baseline differences between the study groups prior to the ablation procedure, as these initial scores are unrelated to the intervention. The number of patients with AF recurrences, re-hospitalizations, or emergency visits was analyzed with the Chi-square statistic (with a Yates correction in case of cell counts <10). Survival free from AF was tested with the log-rank statistic. The Grambsch and Therneau test was significant (p=0.03) demonstrating a violation of the proportionality of hazards assumption, and thereby a Cox regression was not performed. The prevalence of smokers, daily drinkers and patients diagnosed with OSA in the two groups at the 12-month assessment was analyzed with a generalized linear model and a binomial function link, adjusting for the baseline status. The Firths’ correction was used in those analyses with complete separation. Comparisons for patient satisfaction and knowledge on AF (AFKAT questionnaire) were conducted with a linear regression. In the case of non-normal distribution of the residuals, the transformation type was selected based on the lambda value of the boxcox function (MASS package). Statistical significance was set at p≤0.05 for all analyses.

For sample size estimation, we estimated a post-ablation mean ASTA score of 6 and a SD of 3.5 in the Control group. We calculated that 30 patients per group would be needed to detect a mean improvement of 2.5 points in the ASTA score (effect size 0.71) at a significance level (α) of 0.05 and statistical power (1-β) of 0.8. Assuming a 10% loss to follow-up, a total of 66 patients would be needed with a 1:1 randomization ratio. All analyses were conducted with SPSS v26 (IBM, Armonk, NY, USA) and R v4.1 (R Foundation for Statistical Computing, Vienna, Austria).

## Results

The patient flowchart is plotted in **Figure 2**. From March 2022 to March 2023, 116 patients were screened, and 50 patients were excluded, mostly re-do procedures or patients who refused to participate. A total of 66 patients were eventually included and randomized (33 per arm). No patients were lost to follow-up. One patient in the Control group declined having the 6- and 12-month Holter done; he otherwise was asymptomatic and had no AF recurrences. We followed a conservative approach, and the patient was considered as a non-recurrent in the Control group for atrial arrhythmia-free survival analyses. A sensitivity analysis censoring the patient after the 3-month Holter yielded the same conclusions.

**Figure 2.**
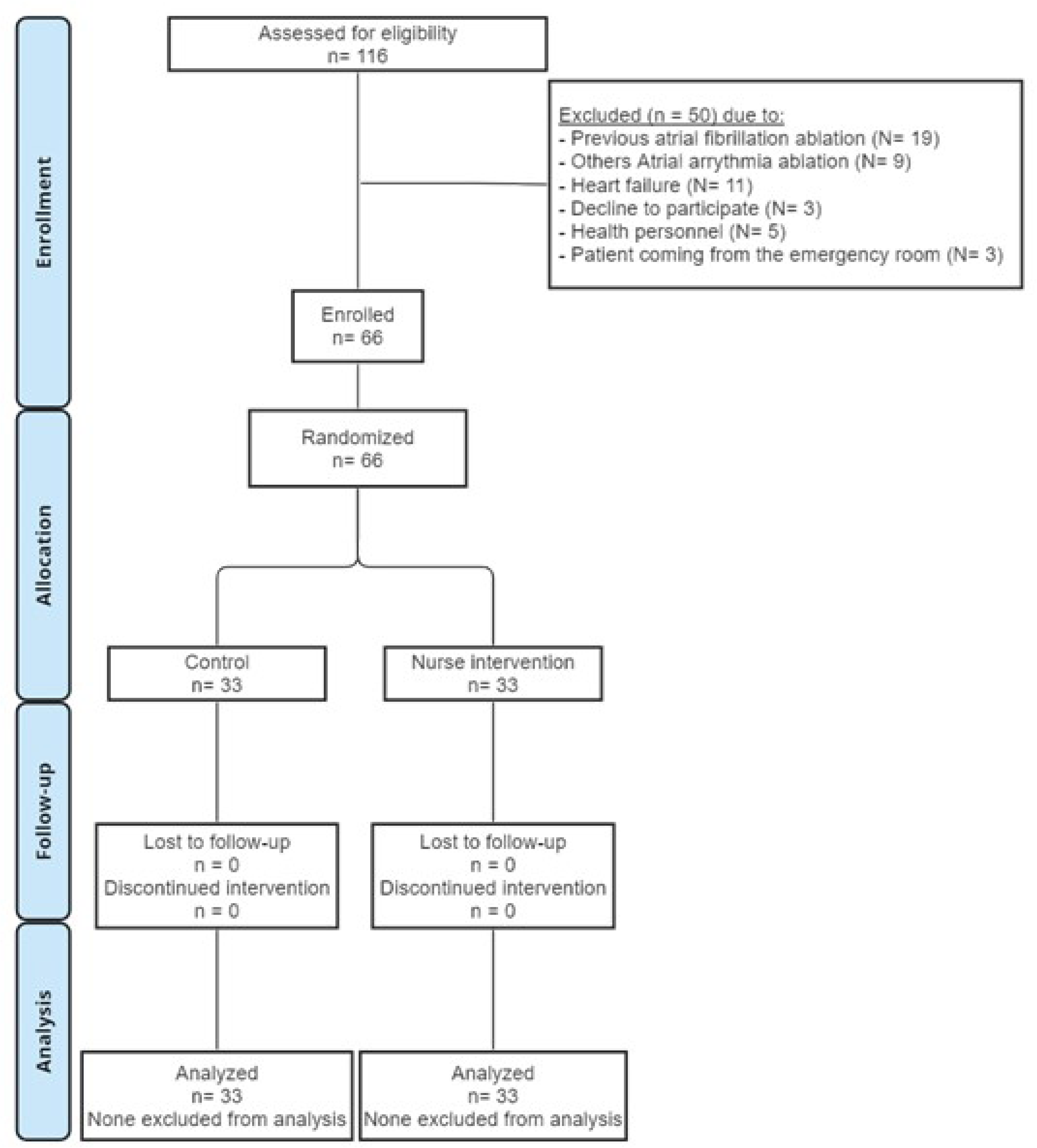
CONSORT diagram of the study design.

Baseline characteristics are shown in **Table 1**. Both experimental groups were well balanced and representative of a general AF ablation cohort. The mean age was 63±10 years, 44 (67%) were men, and two thirds had paroxysmal AF. The preprocedural pharmacological treatment was similar in both groups.

**Table 1.** Characteristics of participants according to treatment group. Data are presented as the mean value ± SD or number (%) of patients;. SD, standard deviation; BMI, body mass index; AF, atrial fibrillation; PAF, paroxysmal atrial fibrillation; OAC, oral anti-coagulants; NOAC, new oral anti-coagulant; VKA, vitamin-K antagonists;

| Characteristics | Control<br>(n=33) | Nurse-led<br>(n=33) |
| --- | --- | --- |
| Age (years) | 63±9.8 | 62±8.4 |
| Male sex, n (%) | 22 (67%) | 23 (70%) |
| AF pattern, n (%) |  |  |
| - Paroxysmal | 20 (61%) | 23 (70%) |
| - Persistent | 13 (39%) | 10 (30%) |
| Hypertension | 23 (70%) | 23 (70%) |
| Diabetes mellitus | 6 (18%) | 4 (12%) |
| Obstructive sleep apnea | 10 (30%) | 7 (21%) |
| Daily drinker | 5 (15%) | 7 (21%) |
| Current smoker | 5 (15%) | 5 (15%) |
| Functional class |  |  |
| - I | 30(91%) | 29 (88%) |
| - II | 3 (9.1%) | 4 (12%) |
| CHA<sub>2</sub>DS<sub>2</sub>-VASc, n (%) |  |  |
| - 0 | 4 (12%) | 2 (6.1%) |
| - 1 | 11 (33%) | 15 (46%) |
| - >2 | 18 (55%) | 16 (49%) |
| LVEF (%) | 58±6 | 56±6% |
| Left atrial diameter (mm) | 39±8 | 41±11 |
| Body mass index (kg/m<sup>2</sup>) | 29.9±3.9 | 29.5±4.7 |
| Antiarrhythmic drugs, n(%) |  |  |
| - Amiodarone | 10 (30%) | 15(46%) |
| - Flecainide | 10(30%) | 11(33%) |
| - Dronedarone | 1 (3%) | 0 |
| - Digoxine | 1(3%) | 0 |
| - Bisoprolol | 18(55%) | 23(70%) |
| - Sotalol | 2(6.1%) | 1(3%) |
| - Class Ic Pill-in-the-pocket | 1 (3%) | 0 |
| Oral anticoagulation, n (%)<sup>6</sup> |  |  |
| - VKA | 2(6.1%) | 4(12%) |
| - DOAC | 31 (94%) | 29 (88%) |

### Quality of life

AF ablation associated with an improvement in QoL in both groups (**Table 2**). For the primary endpoint assessment at 12 months, the QoL measured with the ASTA-QoL questionnaire scored better in the NLC group than in the Control group (0 [IQR:0, 0] in NLC vs. 2 [IQR 0, 7] in usual care, p<0.001, baseline-adjusted estimated difference 4 (95%CI 1.76-6.24), **Figure 3**). Outcomes were comparable in both sexes (P-value for interaction 0.24). Results in the mental and physical domains consistently favoured the NLC group (**Table 2)**. The Cantril ladder also yielded better scores for the NLC than the UC group (8 [7-8] vs 7 [6,8], NLC vs usual care, p=0.02; baseline-adjusted difference 0.93 [95%CI 0.13-1.72]).

**Figure 3.**
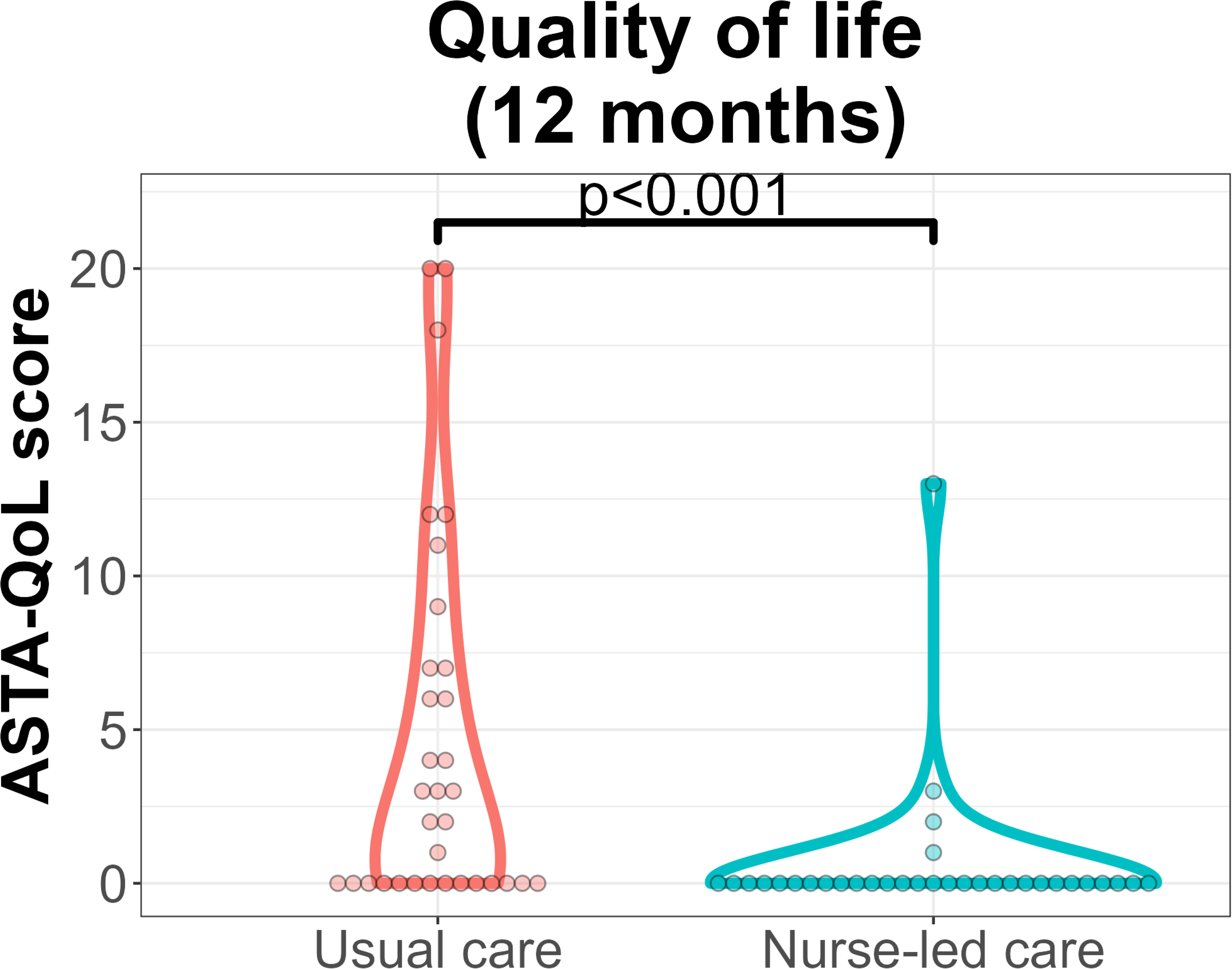
Assessment of the primary endpoint. Violin plot and individual measurements of the ASTA-QoL questionnaire at the 12-month timepoint in both experimental groups. Comparison with long-transformed values and adjusted by baseline values.

**Table 2.** Results of ASTA-QoL questionnaire (mean [interquartile range]) in both groups at baseline and after 12 months to quality-of-life evaluation. “p” reports the baseline-adjusted difference between groups.

|  | Control (n=33) | Intervention (n=33) | p |
| --- | --- | --- | --- |
| **ASTA-QoL score** |  |  |  |
| **Baseline** |  |  |  |
| - Global | **11 (6, 14)** | **10 (5, 16)** |  |
| - Mental | 5 (3, 7) | 5 (2, 7) |  |
| - Physical | 6 (2, 9) | 5 (1, 9) |  |
| **12 months** |  |  |  |
| - Global | **2 (0, 7)** | **0 (0, 0)** | **<0.001** |
| - Mental | 0 (0, 3) | 0 (0, 0) | 0.003 |
| - Physical | 1 (0, 5) | 0 (0, 0) | <0.001 |

### Secondary Endpoints

#### AF recurrences, emergency department visits and unplanned readmissions

NLC significantly reduced the number of patients who presented with AF recurrences compared with the Control group (15 [46%] vs 5 [15%], OR 4.6 [95%CI 1.3-18.9], p=0.014). The survival-free from atrial arrhythmias was consistently higher in the NLC group (**Figure 4A**), similarly in males and females **(Supplemental figure 1).** Of note, recurrences were similar in both groups until 6 months post-ablation, and subsequently progressively diverges. In keeping with these findings, patients in the NLC group required less emergency department visits (19 [58%] vs 7 [21%], Control vs NLC, OR 0.2 [95%CI 0.06-0.66], p=0.006, **Figure 4B**) than patients in the Control group. In the Control group, 1 patient consulted twice and one patient thrice; an analysis with Poisson regression considering the number of visits yielded similar conclusions (OR 0.3 [95%CI 0.12-0.71], p=0.004). Most emergency visits were due to symptomatic AF recurrences (a detailed summary of all reasons for consultation is plotted in **Supplemental figure 2**). Readmissions were numerically higher in the Usual care than in the NLC group, but differences did not reach formal significance (**Figure 4C**)

**Figure 4.**
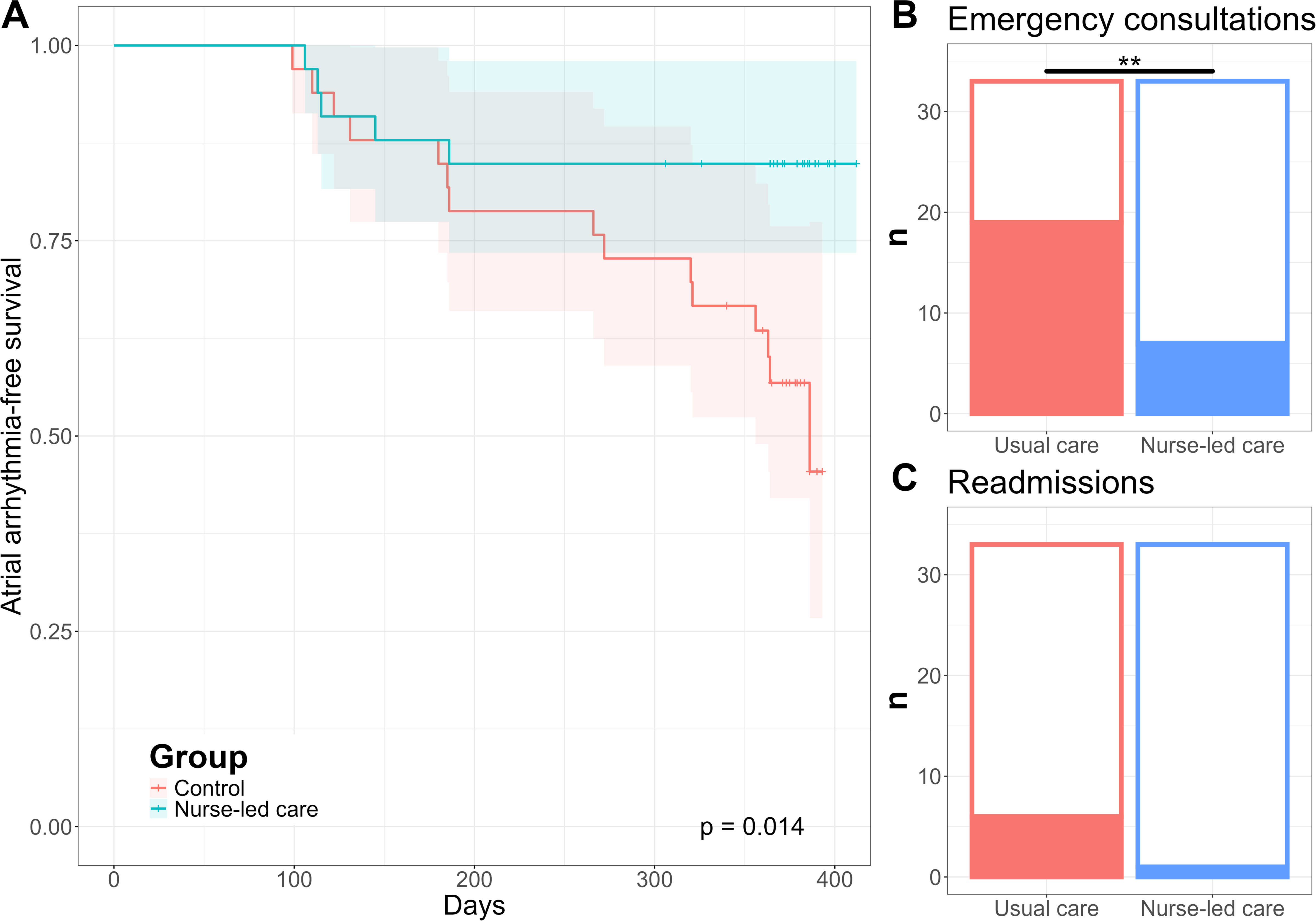
Clinical outcomes in both study groups. (A) Atrial arrhythmia-free survival, (B) visits to the emergency department and (C) unplanned admissions in both groups during the 12-month follow-up. Analysis with a log-rank test (A) and generalized linear models (B, C). * p<0.05, **p<0.001

#### Risk factor assessment

The risk factor burden was similar in both groups at baseline (**Table 1**). During the study period, 13 patients (39%) in the NLC group underwent a PSG based on a high Berlin test, and none in the Control group. Eventually, at 12 months more patients in the NLC group had been diagnosed with severe OSA requiring CPAP therapy (OR 21.4 [95%CI 2.4 – 2835.1], **Figure 5A**). At the end of the experimental period, smoking prevalence was lower in the NLC than in the control group (OR 0.03 [95%CI 0.0002 – 0.55], **Figure 5B**) while daily drinkers were similar in both groups (OR 0.27 [95%CI 0.02 – 2.8], **Figure 5C**). At baseline, most patients were overweight (31 [94%] in the control and 27 [82%] in the NLC groups); 16 patients in the NLC group were referred to an Endocrinologist/Nutritionist for body weight management. Amongst those patients with overweight at baseline, BMI at the 12-month assessment was lower in the NLC than in the control group (estimated difference -0.59 kg/m^2^ [95%CI -1.48 – 0.19], p=0.12, **Figure 5D).** The overall exercise load assessed with the VREM questionnaire was higher in the NLC than in the Control group (estimated difference between groups 1819 METS [95%CI 1105 – 2533], p<0.001, **Figure 5E**). Overall, these results point to an improved control of cardiovascular risk factors in the NLC group.

**Figure 5.**
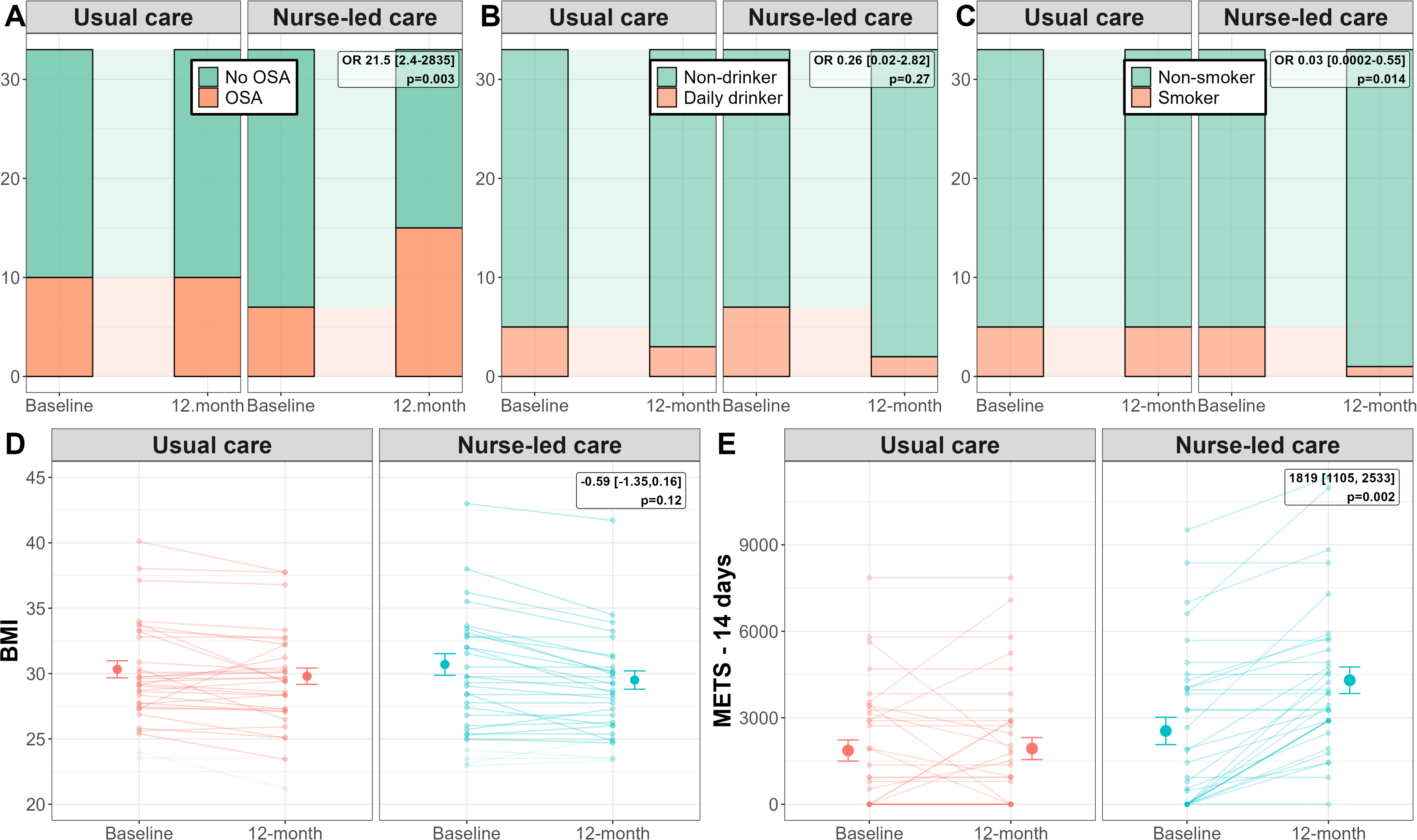
Assessment of AF risk factors at the 12-month timepoint. The prevalence of a diagnosis of obstructive sleep apnea (A), daily drinking (B) and smoking (C), as well as body mass index (D) and physical activity load in METS in the last 14 days is reported in both groups. Analyses with a generalized linear model with a binomial link (A-C) or linear regression models (D, E); in all cases 12-month data was adjusted for the baseline value. The p-value of the group effect is shown in the upper left corner of each panel.

#### Symptomatology

Patients in the NLC group had significantly lower odds of being in a worse mEHRA category at 1 year, compared to usual care, after adjusting for their baseline severity (p=0.016, **Figure 6A**). More patients in the NLC group than in the control group remained asymptomatic (mEHRA 1) (88% vs 61%, p=0.011). Consistently, patients in the NLC group experienced fewer symptoms during the 12 months after ablation as indicated by better ASTA-Symptoms scores (2 [0,7] vs 0 [0,0], estimated baseline-adjusted difference 3.2 [95%CI 1.6 – 4.9], p=0.001, **Figure 6B).** The most frequently reported symptoms in both groups were fatigue, weakness, anxiety, and breathlessness during activity.

**Figure 6.**
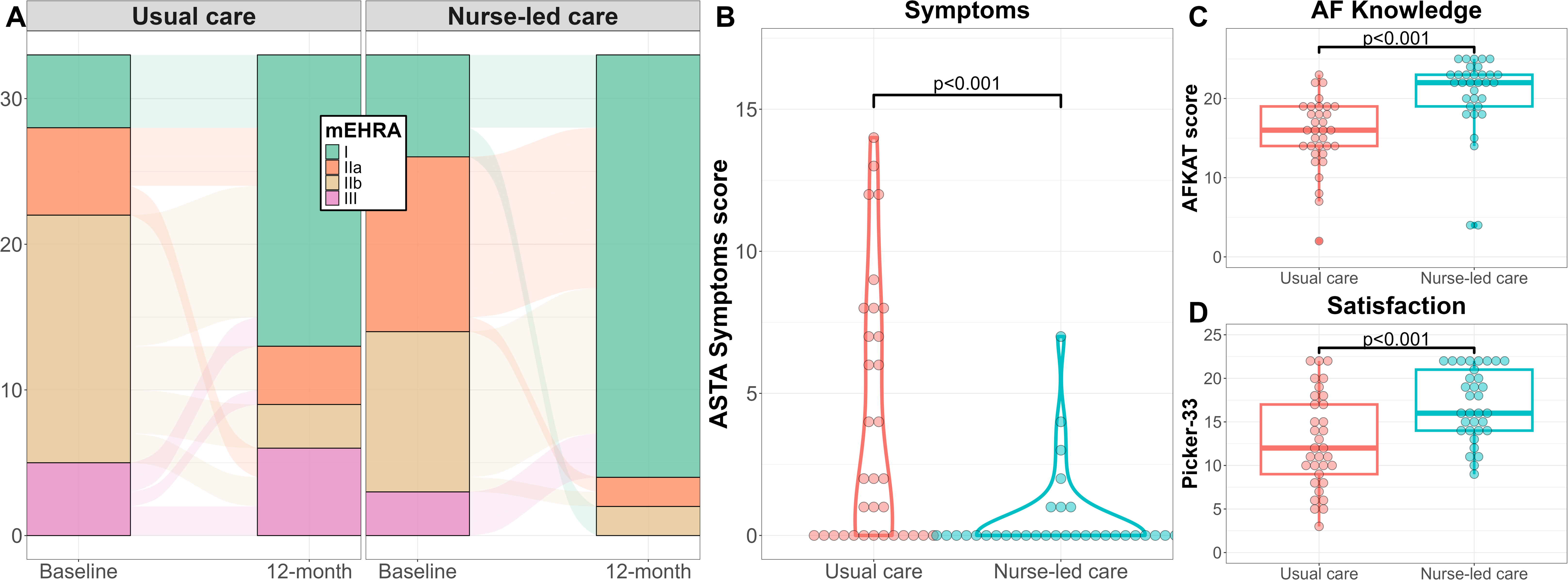
Assessment of secondary endpoint. (A) Symptom assessment with the mEHRA scale at baseline at the 12-month timepoint. (B) Violin plot and individual assessment of quality of life with the ASTA-symptoms questionnaire. (C) Results (boxplot and individual points) of the AFKAT questionnaire assessing knowledge of AF. (D) Results (boxplot and individual points) of the PPEQ-15 questionnaire assessing patients satisfaction of the whole medical process. Comparisons with linear models; in A the baseline value was added as a covariate.

#### Knowledge of AF

The knowledge on AF was assessed in both group at the 12-month visit. Patients in the NLC group demonstrated greater knowledge compared with the control group (20.5±5.1 vs 15.5±4.5, respectively, mean estimated difference 5 [95%CI 3.1, 6.9], p<0.001, **Figure 6C**).

#### Patient satisfaction

Patients in the intervention group reported better patient experience than the control group (17±4.1 vs. 12.7±5.4, estimated difference 4.3 [95%CI 1.9 - 6.6], p<0.001, **Figure 6D**) according to the score obtained in the PPEQ-15.

#### Unplanned contacts

Between March 2022 and March 2023 a log was maintained documenting calls received by the AF nurse practitioner. A total of 25 phone calls were registered and responded to from patients in the NLC group, with 26% occurring prior to ablation and 74% following the procedure. The majority of these calls addressed issues related to pharmacological treatment (12%), complementary tests and follow-up visits (16%), and AF recurrences (16%) or complications (15%).

## Discussion

The NURSECAT-AF randomized controlled trial evidenced that a nurse-led care intervention in patients undergoing AF ablation improved QoL, decreased the risk of recurrences, and the presence of symptoms, and improved the knowledge of patients on AF. This trial introduces an evidence-based, standardized, and scalable nurse-led care model that is initiated prior to catheter ablation and extends into both the short- and long-term follow-up, aiming to optimize clinical outcomes in this patient population.

### Integrated care management and the role of nurse management in atrial fibrillation

AF increases the risk of thromboembolism and heart failure, impairs quality of life, and associates with heightened mortality, thereby justifying strategies aimed at maintaining sinus rhythm.^22^ However, rhythm control in isolation is often insufficient. While conventional pharmacological and interventional therapies remain central to AF management, there is growing recognition of the value of a comprehensive, multidisciplinary approach. This model emphasizes coordinated collaboration among cardiologists and other specialists, primary care physicians, nurses, and allied health professionals, targeting not only rhythm control but also associated comorbidities, patient education, and lifestyle interventions.^23^ In primary care settings, nurse-led integrated care models yield substantial clinical benefits, including improved adherence to guideline-recommended therapies,^24^ enhanced patient satisfaction and QoL,^25^ and even reduced AF-related hospitalizations and cardiovascular death risk.^26,27^

Catheter ablation represents the most effective intervention for maintaining sinus rhythm; however, evidence regarding the potential added value of nurse-centered care in enhancing patient outcomes remains limited. We confirmed here that ablation dramatically improves QoL in most patients with AF. However, despite the superior efficacy of ablation, impaired QoL is still a concern in many patients, especially those with persistent AF or presenting recurrences. Observational studies^28^ and small or short-term trials^29^ support improved outcomes in patients undergoing a nurse follow-up clinic. The NURSECAT-AF trial offers the most robust evidence to date, being the largest and longest randomized controlled trial conducted in the periprocedural context. Our NLC approach was inherently multidimensional, a characteristic likely underpinning its favorable impact on clinical outcomes. Despite being scheduled to undergo an AF-related surgical procedure, patients commonly exhibit substantial knowledge deficits.^30^ Unlike prior studies,^31^ the educational component of the intervention was implemented before ablation, and was complemented with physical informational resources, and a direct interaction with an expert nurse was offered if needed. Patient education and the delivery of high-quality, up-to-date information^32,33^ enhance patient empowerment which in turn supports improved therapeutic adherence -a key determinant in mitigating symptoms and thromboembolic risk associated with AF.^34^ Patients were provided with practical tools to recognize and manage symptoms and recurrences, likely substantiating better symptom control and reduced emergency department visits. However, whether targeted education by itself is enough to reduce hospitalizations is controversial.^35,36^ The promotion of self-awareness of individual risk factors and the benefits of their modification contributed to broader improvements in overall cardiovascular health. Collectively, these factors, alongside a reduction in AF recurrences, eventually culminated in better QoL.

### Improved risk factor management likely mediates a decreased arrhythmic burden

Beyond the observed improvement in quality of life, our findings indicate that a periprocedural NLC programme decreases recurrent AF. Although the primary aim of this study was not to elucidate underlying mechanisms, the observed effects may plausibly be attributed to an improved risk factor profile, aligning with the established benefits of integrated care management on AF recurrences. Notably, the nurse intervention in the NLC group enabled uncovering a substantial number of previously undiagnosed OSA cases, whereas no new diagnoses were made in the control group throughout the study period. While the benefits of continuous positive airway pressure on AF risk remain debated,^37,38^ increased diagnostic recognition may have prompted a broader focus on risk factor control, and continuous positive airway pressure contributed to improve QoL. Additionally, patients in the NLC group demonstrated a significantly higher rate of smoking cessation^39,40^ and greater engagement in physical activity.^41^ Among individuals with obesity, those in the NLC arm achieved more substantial weight reduction compared to usual care.^42^ These lifestyle modifications are supported by robust evidence linking them to reduced AF burden. Observational^43^ and large randomized^44^ studies substantiate the benefits of an intensive, multidisciplinary risk factor management in reducing AF recurrences. Importantly, differences in AF recurrence rates became apparent only after six months post-randomization. It is reasonable to speculate that some months are needed to reach an optimal control of weight and physical activity, as well as to complete the diagnostic work-up for OSA. This observation also underscores the potential advantage of initiating risk factor management earlier in the clinical course, suggesting that pre-ablation intervention could confer even greater benefit.

In routine clinical practice, risk factor control is often suboptimal. Several factors may account for the success of the NLC in promoting risk factor modification and reducing AF recurrence in NURSECAT-AF. Patients undergoing AF ablation may be particularly receptive to interventions aimed at disease resolution, and the periprocedural window likely represents an optimal, sweet-spot, period for initiating lifestyle changes. The intervention itself—comprising four structured nurse-led interactions and a direct contact channel providing prompt contact—allowed for robust reinforcement. While the long-term sustainability of these benefits was not assessed, continued reinforcement by primary care professionals may be necessary to maintain favorable outcomes over time.

### Limitations

Some limitations warrant consideration. NURSECAT-AF was conducted as a single-center trial in a tertiary care setting; therefore, additional studies are necessary to validate the generalizability of our results. Unfortunately, data on the management of other comorbidities influencing AF risk, such as hypertension, were not systematically collected. It should be noted, however, that strict blood pressure management in the periprocedural window already failed to decrease AF recurrences in a randomized trial.^45^ Patients in the control group were aware of the existence of the NLC and the closer follow-up of risk factors implemented in the intervention group, introducing a potential a contamination bias that may have attenuated the impact of the NLC. Conversely, social desirability bias could have exaggerated perceived differences in certain parameters; nonetheless, the consistency of objective measures such as BMI or AF recurrences support the robustness of our conclusions.

## Conclusions

Nurse-led integrated care for patients undergoing AF ablation yields substantial improvements in quality of life, symptomatic relief and reduces arrhythmia recurrences, readmissions, and emergency department visits at one-year post-procedure.

## Non-standard Abbreviations and Acronyms

AF: Atrial Fibrillation
ASTA: Arrhythmia-Specific Scale in Tachycardia and Arrhythmia
NLC: Nurse-led care
QoL: Quality of life
OSA: Obstructive Sleep Apnoea
PSG: Polysomnography
BMI: Body Mass Index
EHRA: European Heart Rhythm Association
CPAP: Continuous Positive Airway Pressure

## Acknowledgments

We are indebted to the nurse team, clinical and research fellows for their contribution to this trial, and Mrs. Eulalia Ventura, Mrs. Carol Sanroman, Mrs. Jessica Soriano and Mr. Jose Alcolea for their administrative support. We would like to thank Núria Fabrellas and Adela Zabalegui for their leadership in nursing research and their support for our team. We would like to thank Arnau Miguel for his advice as a graphic designer.

## Sources of Fundings

This study has received funding for Col·legi Oficial d’Infermeria de Barcelona (COIB) [PR-647/2023]; Instituto de investigación Sanitaria Valdecilla (IDIVAL) [ENFVAL23/07]; and Instituto de Salud Carlos III [PI22/00953]. ACV was awarded with a “Beca d’intensificació La Pedrera 2021” which provided temporary full-time dedication to the trial.

## Disclosures

The authors declare no conflicts of interest relevant to the topic of this study.

## Authors’ Contributions

ACV designed, conceptualized and oversaw the project, contributed to patient inclusion and follow-up, recorded follow-up data, contributed to data interpretation, and wrote the first draft of the manuscript. LM contributed to study design and conceptualization, and critically reviewed the final draft. EG contributed to study design and conceptualization, performed statistical analyses, interpretated the results, and critically reviewed the first and final draft. MAMM: contributed to study conceptualization and critically reviewed the final draft. ECF, MNB, RCD, SHP, MVN contributed to patient inclusion and critically reviewed the final manuscript. RB contributed to statistical analyses of the data. JMT, APS, JBG, TA, IRL contributed to patients follow-up, interpretation of the results and critically reviewed the final manuscript.

## Data availability

Raw data may be made available from the corresponding authors upon reasonable request. Supplemental publication material for this article is available online.

## Notes

### Competing Interest Statement

The authors have declared no competing interest.

### Clinical Trial

NCT05333445

### Funding Statement

This study has received funding for Collegi Oficial d'Infermeria de Barcelona (COIB) [PR-647/2023]; Instituto de investigación Sanitaria Valdecilla (IDIVAL) [ENFVAL23/07]; and Instituto de Salud Carlos III [PI22/00953]. ACV was awarded with a Beca d'intensificació La Pedrera 2021 which provided temporary full-time dedication to the trial.

### Author Declarations

OPINION OF THE ETHICS COMMITTEE ON MEDICINE RESEARCH ANA LUCIA ARELLANO ANDRINO, Secretary of the Ethics Committee for Research with Medicines at the Hospital Clinic de Barcelona (HCB), Certificate: That this Committee has evaluated the promoter's proposal, so that the study can be carried out: COde: Reg. HCB/2021/1045 VERSION: V2.13.10.2021 TITLE: Randomized controlled study on the effect of the educational intervention nurse-led ongoing to atrial fibrillation ablation PROMOTER: Hospital Clinic de Barcelona, Fundacio Clinic - IDIBAPS, universitat de Barcelona. PRINCIPAL INVESTIGATOR: ALBA CANO VALLS and considers that, taking into account the response to the clarifications requested (if any), and that: - The necessary requirements for the suitability of the protocol in relation to the objectives of the study are met and the foreseeable risks and inconveniences are justified. - The researcher's capacity and the available resources are appropriate for carrying out the study. - The planned financial compensation (if any) and its possible interference with ethical principles have been evaluated and are considered appropriate. - The study complies with the essential ethical standards and deontological criteria governing this centre. - The study complies with the obligations established by the applicable research and confidentiality regulations. - That the study is included in one of the accredited lines of biomedical research at this centre, meeting the necessary requirements, and that it is viable in all respects. This CEIm accepts that the study may be carried out, and any changes to the protocol or serious adverse events must be reported to the Ethics Committee. and certifies that: 1. At the meeting held on 7 October 2021, minutes 17/2021, it was decided to issue the report corresponding to the reference study. 2. The CEIm of the Hospital Clinic i Provincial, both in its composition and in its PNTs, complies with the standards of EMA/CHMP/ICH/135/1995. 3. List of members: Chair: - JOAQUIM FOReS I VInETA (Traumatologist, HCB) Vice-President: - JOSEP MARiA MIRo MEDA (Infectious Diseases Specialist, HCB) Secretary: - ANA LUCIA ARELLANO ANDRINO (Clinical Pharmacologist, HCB) Members: - JOSE RIOS GUILLERMO (Statistician. Medical Statistics Platform. IDIBAPS) - OCTAVI SANCHEZ LOPEZ (Patient representative) - MARIA JESuS BERTRAN LUENGO (Epidemiologist, HCB) - JOAQUiN SaEZ PEnATARO (Clinical Pharmacologist, HCB) - SERGI AMARO DELGADO (Neurologist, HCB) - EDUARD GUASCH CASANY (Cardiologist, HCB) - MARINA ROVIRA ILLAMOLA (Primary Care Pharmacist, CAP Eixample) - PAU ALCUBILLA PRATS (Clinical Pharmacologist, HCB) - JOSE TOMAS ORTIZ PEREZ (Cardiologist, HCB) - ELENA CALVO CIDONCHA (Hospital Pharmacist, HCB) - CECILIA CUZCO CABELLOS (Nurse, HCB) - PAULA MARTiN FARGAS (Lawyer, HCB) - SALVATORE BRUGALETTA (Cardiologist, HCB. Member of the CEA, HCB) - XAVIER CANALS-RIERA (Telecommunications Engineer) - FRANCESC XAVIER CORBELLE (Computer Scientist, HCB) - JOSEP DiAZ CORT (Graduate in Physical Sciences. Professor of Computer Science) - GASPAR MESTRES ALOMAR (Doctor, Angiology, Vascular Surgery, HCB) - MARTA FRANCH SAGUER (Lawyer) - PATRICIA AMOROS REBOREDO (Hospital Pharmacist, HCB) - ANNA MARiA GUIJARRO PeREZ (Citizen Services, HCB) - JULIO DELGADO GONZaLEZ (Hematologist, HCB) Barcelona, a 13 de diciembre de 2021

